# Clinical features and outcomes of hospitalised patients with COVID-19 and Parkinsonian disorders: a multicentre UK-based study

**DOI:** 10.1101/2023.04.24.23289022

**Authors:** Lexy Sorrell, Valentina Leta, Anton Barnett, Kara Stevens, Angela King, Jemma Inches, Christopher Kobylecki, Richard Walker, K Ray Chaudhuri, Hannah Martin, Jane Rideout, J Robert Sneyd, Sarah Campbell, Camille Carroll, the COVID-19 PD UK study team

## Abstract

**Background:** Parkinson’s disease has been identified as a risk factor for severe Coronavirus disease 2019 (COVID-19) outcomes. However, whether the significant high risk of death from COVID-19 in people with Parkinson’s disease is specific to the disease itself or driven by other concomitant and known risk factors such as comorbidities, age, and frailty remains unclear.

**Objective:** To investigate clinical profiles and outcomes of people with Parkinson’s disease and atypical parkinsonian syndromes who tested positive for COVID-19 in the hospital setting in a multicentre UK-based study.

**Methods:** A retrospective cohort study of Parkinson’s disease patients with a positive COVID-19 test admitted to hospital between February 2020 and July 2021. An online survey was used to collect data from clinical care records, recording patient, Parkinson’s disease and COVID-19 characteristics. Associations with time-to-mortality and severe outcomes were analysed using either the Cox proportional hazards model or logistic regression models, as appropriate.

**Results:** Data from 552 admissions were collected: 365 (66%) male; median (inter-quartile range) age 80 (74-85) years. The 34-day mortality rate was 38.4%; male sex, increased age and frailty, Parkinson’s dementia syndrome, requirement for respiratory support and no vaccination were associated with increased mortality risk. Community-acquired COVID-19 and co-morbid chronic neurological disorder were associated with increased odds of requiring respiratory support. Hospital-acquired COVID-19 and delirium were associated with requiring an increase in care level post-discharge.

**Conclusions:** This first, multicentre, UK-based study on people with Parkinson’s disease or atypical parkinsonian syndromes, hospitalised with COVID-19, adds and expands previous findings on clinical profiles and outcomes in this population.

## Introduction

The Coronavirus disease 2019 (COVID-19) pandemic caused by severe acute respiratory syndrome coronavirus 2 (SARS-CoV-2) has spread worldwide since early 2020 with unprecedented speed (1). Clinical risk prediction models have been developed to identify risks of poor short-term outcomes due to the SARS-CoV-2 infection (mortality and hospital admission) and offer guidance to public health policymakers for clinical decision-making processes, prioritisation for vaccination, and targeted recruitment for clinical trials. Among them, the QCovid and the more recently developed QCovid3 risk algorithms, have identified Parkinson’s disease (PD) as a risk factor for severe COVID-19 outcomes, despite vaccination (adjusted cause-specific hazard ratios for COVID-19 death after vaccination of 2.23 (1.79 to 2.78)) (2, 3). Advanced age, frailty and impaired cough reflex are commonly observed in people with PD (PwP) and might contribute to their susceptibility to developing severe acute respiratory syndrome (4). A variety of studies have investigated mortality in PwP and COVID-19 with figures ranging from 5% to 100% (5); the wide range reflects the heterogeneity of methodologies used (case report, series, surveys, retrospective or prospective cohort studies) and cohort analysed (home-based vs hospitalised patients, early vs advanced patients, etc.) (6–18). However, whether the significant high risk of death from COVID-19 in PwP is specific to PD itself or driven by other concomitant and known risk factors such as comorbidities (hypertension, diabetes, pulmonary disease, obesity, immunosuppression, and dementia), older age (>60 years), and frailty remains unclear (5). The COVID-19 PD study is the first to investigate the association of demographic, co-morbidity, COVID-19 and PD-specific factors with mortality and severe outcomes of inpatients with PD and atypical parkinsonian syndromes (APS) who tested positive for COVID-19 admitted to a UK NHS trust hospital.

## Methods

### Study design and population

The COVID-PD UK study was a retrospective cohort study across 21 acute care settings in England funded by Parkinson’s UK (Ref: G-2001). Approvals were obtained from the University of Plymouth Faculty Research Ethics and Integrity Committee (Ref: 2269) and the Health Research Authority (IRAS ID 285686). Members of the project team included PwP who were involved in all aspects of protocol and study development.

The study consisted of patients with a clinical diagnosis of PD, APS (including progressive supranuclear palsy, multiple system atrophy) or Parkinson’s dementia syndrome (Parkinson’s disease dementia (PDD) or dementia with Lewy bodies (DLB)) admitted to participating hospitals between 5^th^ February 2020 and 31^st^ July 2021, with a positive polymerase chain reaction (PCR) test. Exclusion criteria were patients with a diagnosis of vascular parkinsonism and a COVID-19 positive test over 2 weeks prior to admission or at any time following discharge.

Clinical care teams completed an online survey on JISC (https://www.onlinesurveys.ac.uk/) using patients’ clinical care records, extracting information from admission to at least 28-days following admission. Retrospective data collection began in February 2021 and the online survey closed on the 31^st^ July 2021. Individuals were pseudo-anonymised by sites to allow for data clarification where required.

Data included patient, PD and COVID-19 related characteristics along with details of admission, discharge and participation in a COVID-19 related clinical trial. Comorbidities were chosen to allow comparison with the International Severe Acute Respiratory and emerging Infection Consortium (ISARIC) study of hospitalised COVID-19 patients in the UK (19). Patient characteristics included age, sex, ethnicity, index of multiple deprivation (IMD), location pre-admission and clinical frailty score (CFS) (20). PD related features captured included those considered neurological risk features in UK national guidance (21), such as significant cognitive impairment or psychosis, bulbar symptoms, significant respiratory compromise, significant autonomic neuropathy, as well as marked motor fluctuations and Hoehn and Yahr (H&Y) stage. PD features and comorbidities could be entered as unknown by site staff on the JISC survey. No imputation of missing data was performed, however, sites were asked to address unexpected responses and missing data during the querying process.

The wave of positive COVID-19 test was identified. Wave one: 23/03/2020 to 30/05/2020, corresponding to wild-type SARS-CoV2, and wave two: 07/09/2020 to 22/05/2021 (22) corresponding to the emergence of alpha and delta variants in the (23).

Patients were classified as vaccinated if their positive COVID-19 test was over 14 days following their first vaccine dose. COVID-19 was classified as hospital-acquired if the positive COVID-19 test was more than 5 days following admission; otherwise, COVID-19 was classified as community-acquired.

COVID-19 and non-COVID-19 symptoms at admission were recorded as free text and subsequently grouped by the research team into categories including altered mental state (delirium, confusion, obtundation, reduced oral intake), COVID-19 (pyrexia, cough, anosmia) and other respiratory symptoms. Symptoms were summarised for the community- and hospital-acquired COVID-19 patients separately.

The primary outcome of the study was death within 28-days of a COVID-19 positive test. Date of death was recorded as the Sunday following death to preserve anonymity. We therefore used death within 34-days as a proxy outcome; this assumption was explored in a sensitivity analysis. Secondary outcomes included the requirement for respiratory support, an increase in the level of care post-discharge and change in levodopa equivalent daily dose (LEDD).

### Statistical methods

A summary of the data is presented using frequency and percentages for categorical variables of the non-missing sample, while means, standard deviations, medians and ranges are presented for continuous variables. Values entered as ‘unknown’ were coded to missing to ease the comparison between presence and absence of characteristics and due to the small group sizes of some unknown categories.

Survival within 34-days of a COVID-19 positive test was analysed using univariable and multivariable Cox proportional hazards models, using Schoenfeld’s test to assess the assumption of proportional hazards. The requirement for respiratory support and an increase in care were modelled using univariable and multivariable logistic regression models. Model selection for the final multivariable models was conducted using backward elimination, whilst including variables of clinical interest: sex, age, ethnicity, diagnosis, wave of COVID-19 positive test and where COVID-19 was acquired, regardless of their statistical significance. For each outcome, the first-order interaction of wave and where COVID-19 was acquired was examined for statistical significance. In the multivariable models, site was included as a random effect to account for potential differences in practice and COVID-19 severity between UK regions during the pandemic.

Change in LEDD was found to occur in less than 25% of discharged individuals and was therefore analysed descriptively.

Sensitivity analyses were conducted for all outcomes using individuals with a positive COVID-19 test in wave two, sites that comprehensively selected their patients to enter over waves one and two and for the survival outcome, censoring at 28-days.

All analyses are presented with 95% confidence intervals and all tests are two-sided, where a p-value of < 0.05 is considered statistically significant. All analyses were conducted using R version 4.1.3 (24) using packages including survival (25), glmmTMB (26) and forester (27).

## Results

Site staff from 21 hospitals entered 627 individual data sets; of these 17 were duplicates, 38 were excluded due to unverifiable data and 20 were excluded due to meeting the exclusion criteria. The final number of individual admission data sets included in the COVID-19 PD study was 552 compromising 385 community-acquired and 167 hospital-acquired COVID-19 infection episodes; the timing of these admissions is shown in Figure 1. Six sites entered data comprehensively over waves one and two (n=242), while due to resource constraints, the other sites submitted data for one of the two waves or entered data from a selection of patients.

**Figure 1:**
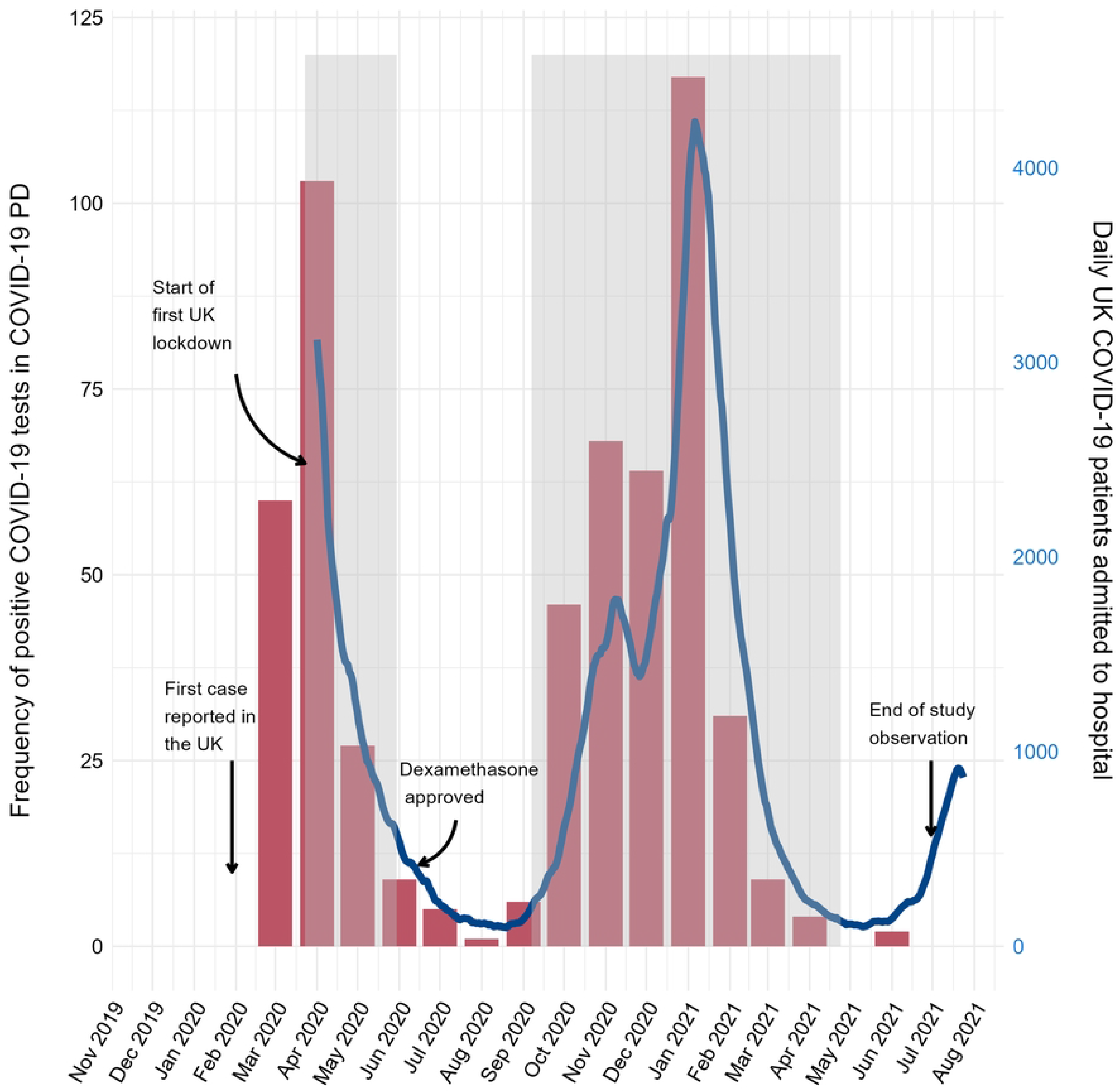
Positive COVID-19 cases captured in the COVID-19 PD study (red bar chart, left y-axis) by month, overlaid by UK daily COVID-19 hospital admissions (28) (blue line graph, right y-axis). Grey regions reflect COVID-19 wave one (23/03/2020 – 30/05/2020) and wave two (07/09/2020 – 22/05/2021) in the UK.

### Demographics and comorbidities

Patient demographics and diagnosis of parkinsonian syndrome details are presented in Table 1. The majority of the patients had PD (349/552 (63.2%)), with 170/552 (30.8%) having PDD/DLB). Only 33 (6.0%) of patients had APS. The median (interquartile range (IQR)) age at admission was 80 (74-85) years with 73.6% of patients over the age of 75 years; APS patients were younger (median (IQR) 75 (70-78) years), while 365/552 (66.1%) were male. The majority of patients were White British (476/552 (86.2%)); further details of the sample ethnicity can be found in Table S2.1 (see S2 Supporting information). Thirty-two patients were enrolled into COVID-19 therapeutic clinical trials, including 21 in the RECOVERY trial (29). Further information on Parkinson’s and COVID-19 characteristics is provided in Tables S2.1 to S2.3 (see S2 Supporting information).

**Table 1:** Patient demographics by diagnosis of Parkinson’s disease (n=349), Parkinson’s dementia syndrome (n=170) and Atypical Parkinsonian syndrome (n=33). For further breakdown of ethnicity, see Table S2.1 (see S2 Supporting information). Abbreviations: Levodopa equivalent daily dose (LEDD), Index of multiple deprivation (IMD).

|  |  | Parkinson's<br>disease | Parkinson's<br>dementia<br>syndrome | Atypical<br>parkinsonian<br>syndrome | Total |
| --- | --- | --- | --- | --- | --- |
|  | N (% of total) | 349 (63.2) | 170 (30.8) | 33 (6.0) | 552 (100.0) |
| **Age at admission (years)** | N | 349 | 170 | 33 | 552 |
|  | Median (IQR) | 81 (74-86) | 81 (76-85) | 75 (70-78) | 80 (74-85) |
|  | [Min., Max.] | [43, 99] | [65, 101] | [45, 98] | [43, 101] |
| **Disease duration (years)** | N | 339 | 168 | 33 | 540 |
|  | Median (IQR) | 5 (3-9) | 5 (3-9) | 2 (1-4) | 5 (2-9) |
|  | [Min., Max.] | [0, 27] | [0, 29] | [0, 11] | [0, 29] |
| **LEDD (mg)** | N | 340 | 169 | 33 | 542 |
|  | Median (IQR) | 450 (300-653) | 375 (150-575) | 200 (0-400) | 400 (243-615) |
|  | [Min., Max.] | [0, 2313] | [0, 2300] | [0, 800] | [0, 2313] |
| **Sex** | N | 349 | 170 | 33 | 552 |
| Male |  | 224 (64.2) | 120 (70.6) | 21 (63.6) | 365 (66.1) |
| Female | N (%) | 125 (35.8) | 50 (29.4) | 12 (36.4) | 187 (33.9) |
| **Ethnicity** | N | 349 | 170 | 33 | 552 |
| White British |  | 298 (85.4) | 152 (89.4) | 26 (78.8) | 476 (86.2) |
| Asian/British Asian (Indian) |  | 10 (2.9) | 3 (1.8) | 2 (6.1) | 15 (2.7) |
| White (any other background) | N (%) | 10 (2.9) | 2 (1.2) | 1 (3.0) | 13 (2.4) |
| Asian/British Asian (Pakistani) |  | 5 (1.4) | 1 (0.6) | 0 (0.0) | 6 (1.1) |
| Other |  | 26 (7.4) | 12 (7.1) | 4 (12.0) | 42 (7.6) |
| **Hoehn and Yahr** | N | 322 | 167 | 32 | 521 |
| 1-2 |  | 49 (15.2) | 5 (3.0) | 0 (0.0) | 54 (10.4) |
| 2.5-3 | N (%) | 125 (38.8) | 41 (24.6) | 8 (25.0) | 174 (33.4) |
| 4-5 |  | 148 (46.0) | 121 (72.5) | 24 (75.0) | 293 (56.2) |
| **Clinical frailty score** | N | 345 | 167 | 33 | 545 |
| <5 |  | 75 (21.6) | 3 (1.8) | 3 (9.1) | 81 (14.8) |
| 5-6 | N (%) | 170 (49.3) | 71 (42.5) | 14 (42.4) | 255 (46.8) |
| 7-9 |  | 100 (29.0) | 93 (55.7) | 16 (48.5) | 209 (38.3) |
| **Location pre-admission** | N | 349 | 170 | 33 | 552 |
| Local/community hospital |  | 3 (0.9) | 3 (1.8) | 1 (3.0) | 7 (1.3) |
| Own Home/private residence | N (%) | 268 (76.8) | 97 (57.1) | 24 (72.7) | 389 (70.5) |
| Residential or nursing home |  | 78 (22.3) | 70 (41.2) | 8 (24.2) | 156 (28.3) |
| **IMD decile** | N | 349 | 170 | 33 | 552 |
| 1-2 |  | 63 (18.1) | 28 (16.5) | 5 (15.2) | 96 (17.4) |
| 3-4 |  | 77 (22.1) | 23 (13.5) | 4 (12.1) | 104 (18.8) |
| 5-6 | N (%) | 77 (22.1) | 37 (21.8) | 9 (27.3) | 123 (22.3) |
| 7-8 |  | 69 (19.8) | 38 (22.4) | 8 (24.2) | 115 (20.8) |
| 9-10 |  | 63 (18.1) | 44 (25.9) | 7 (21.2) | 114 (20.7) |

More patients hospitalised with positive COVID-19 tests had community-acquired rather than hospital-acquired COVID-19 in both wave one (153/190 (80.5%)) and wave two (220/343 (64.1%)).

509/552 (92.2%) had at least one reported co-morbidity at admission, the most frequently reported being hypertension (251/542, 46.3%), dementia (218/535 (40.7%)), chronic cardiac disease (193/539 (35.8%)) and chronic kidney disease (117/535 (21.9%)) (see Table S2.1, S2 Supporting information).

### Parkinsonian syndrome-related features

349/552 (63.2%) patients had PD, 170/552 (30.8%) had PDD/DLB and 33/552 (6.0%) patients had an atypical parkinsonian syndrome. Overall, 251/535 (46.9%) of the cohort had significant cognitive impairment and 154/493 (31.2%) had marked motor fluctuations. Bulbar symptoms were most common in APS patients (16/32 (50.0%) vs. 49/331, (14.8%) and 33/160 (20.6%) for PD and PDD/DLB, respectively) (see Figure 2 and Table S2.2, S2 Supporting information). Patients with PDD/DLB and APS more often had pre-morbid Hoehn and Yahr stages 4-5 (121/167 (72.5%) and 24/32 (75.0%), respectively) compared to PD patients (148/322 (46.0%).

**Figure 2:**
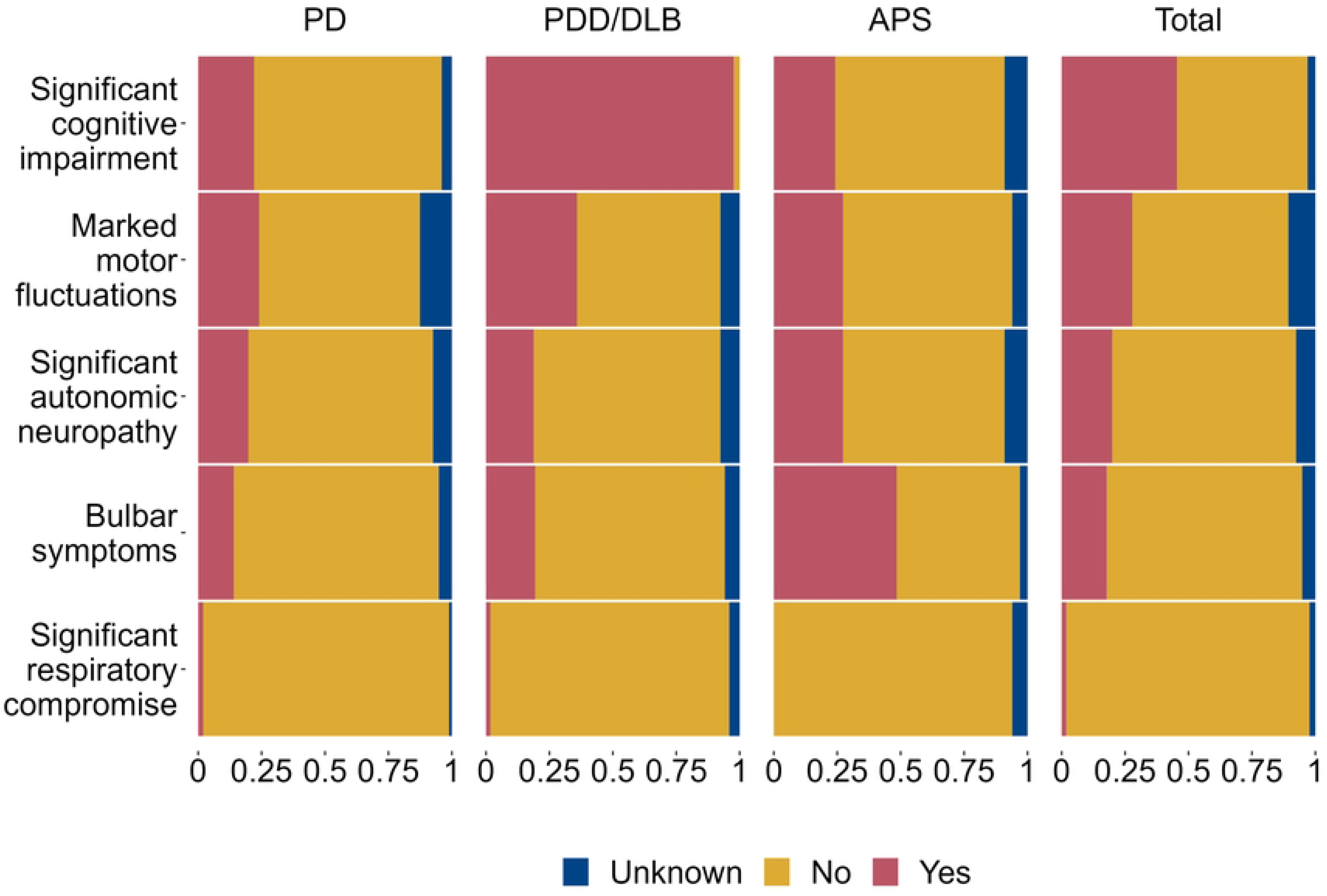
Clinical features of Parkinson’s as a proportion by diagnosis of Parkinson’s disease. X-axis: proportion of feature within diagnosis. Abbreviations: Parkinson’s disease (PD), Parkinson’s dementia syndrome (PDD/DLB) and atypical parkinsonian syndrome (APS).

### Symptoms of COVID-19

The frequency of altered mental state and classical COVID-19 or respiratory symptoms for community- and hospital-acquired COVID-19 patients are presented in Figures 3A and 3B, respectively. Of patients with community-acquired COVID-19, 7.0% (27/385) had neither altered mental state nor classical COVID-19 or respiratory symptoms. This was the case in 25.1% (42/167) of the hospital-acquired COVID-19 patients. Altered mental state was the only feature of COVID-19 infection in 15/385 (3.9%) of community-acquired and 16/167 (9.6%) of hospital-acquired cases.

**Figure 3:**
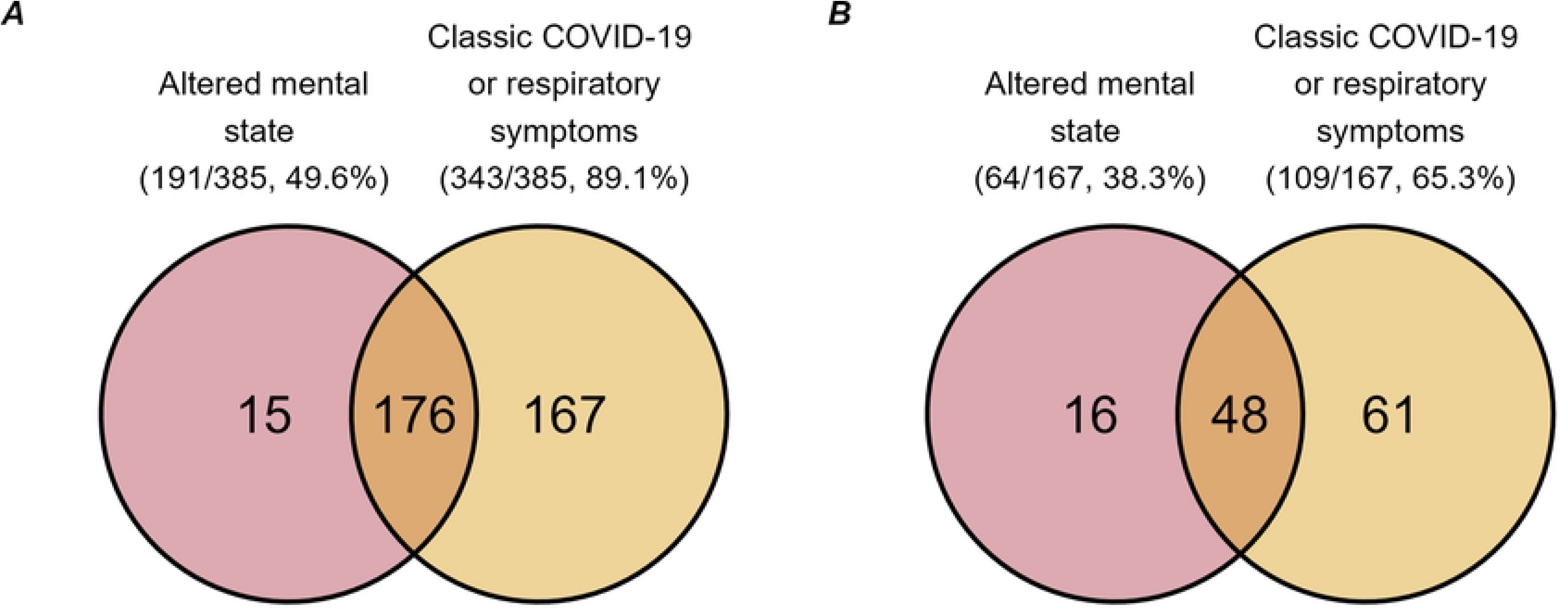
Frequency of altered mental state and classic COVID-19 or respiratory symptoms by where COVID-19 was acquired. A: The frequency of community acquired COVID-19 patients (total n=385) by symptoms of altered mental state, classic COVID-19 or other respiratory symptoms during admission). B: The frequency of hospital acquired COVID-19 patients (total n=167) with the symptoms of altered mental state and classic COVID-19 or respiratory symptoms (mild symptoms or respiratory support) associated with their COVID-19 infection.

### Length of stay

The time from positive test to discharge of patients who acquired COVID-19 in the community was shorter compared to those who acquired COVID-19 in hospital **Error! Reference source not found.**(median (IQR) of 12 (7–21) and 16 (5–28) days, respectively). For those discharged, 84% of community-acquired patients were discharged by day 28 compared to 75% of hospital-acquired patients (Figure 4). Time until discharge was longer for those with bulbar symptoms (median (IQR) of 17 (8–27) compared to 13 (6–23)), a chronic neurological disorder (19 (10–31) compared to 12 (6–22)), those in wave two compared to wave one (15 (6–27) and 12 (7–19), respectively) and those with delirium (15 (7–29) compared to 12 (6–21)). Time until discharge for further patient, COVID-19 and PD characteristics are detailed in Table S2.4 (see S2 Supporting information).

**Figure 4:**
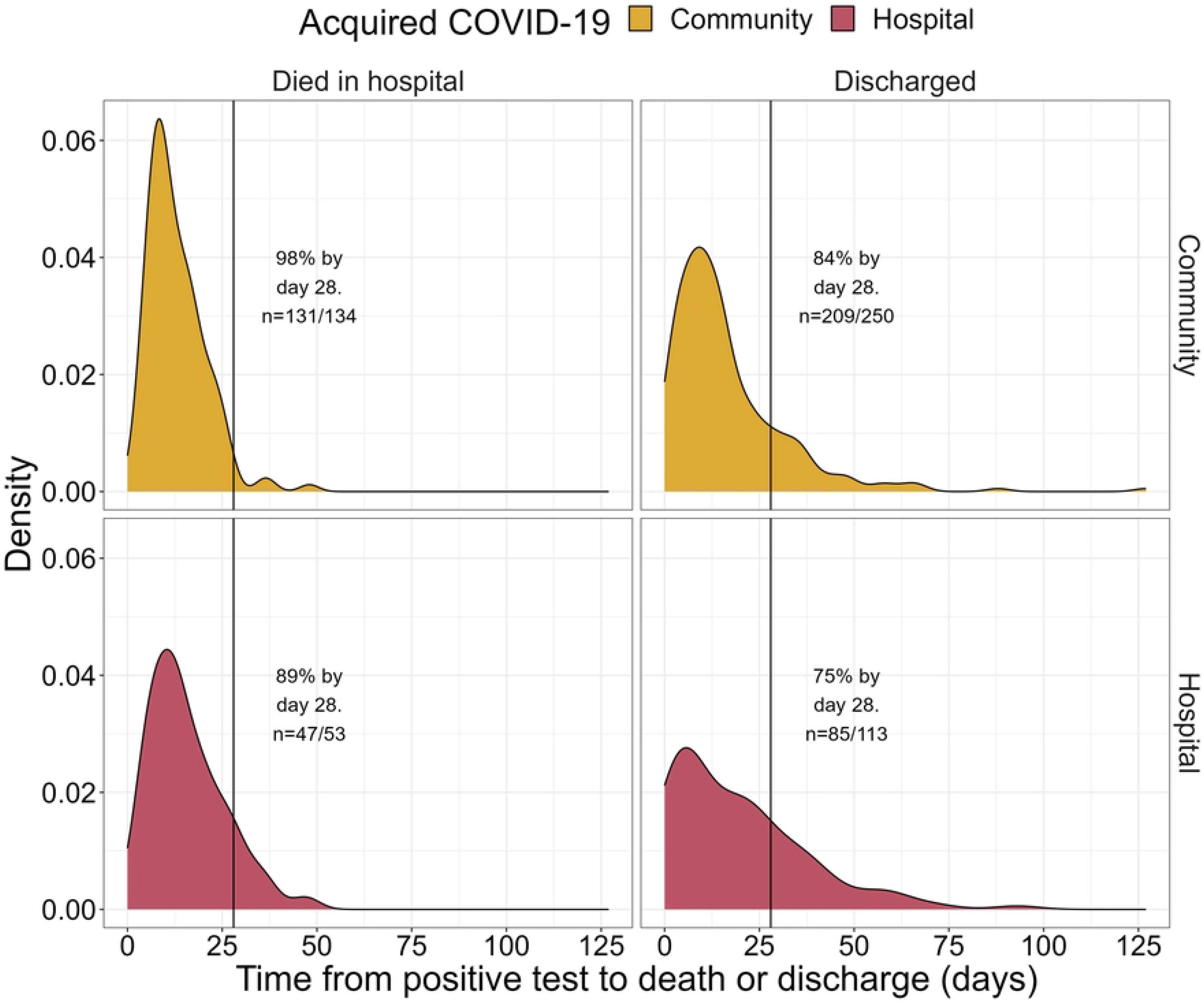
Density plot of time to discharge or death from first positive COVID-19 test by place of COVID-19 acquisition (community or hospital). Two patients that remained in hospital following the study period have been omitted.

### Outcomes

#### Mortality

Details of patient outcomes and discharge are provided in Table S2.5 (see S2 Supporting information). Of the 552 patients in the COVID-19 PD study, 212 (38.4%) died within 34-days of a COVID-19 positive test. Of these patients, 85.4% (181/212) died in hospital, 9.0% (19/212) died following discharge and 5.7% (12/212) died having been discharged to end-of-life care.

Univariable and multivariable results of the Cox models for mortality are provided in Table S3.1 (see S3 Supporting information), along with the results from each sensitivity analysis. The results of the multivariable Cox proportional hazards model are presented in a forest plot in Figure 5. Increased risk of mortality within 34-days was associated with increased age (hazard ratio (HR) 1.05, with a 95% confidence interval (CI) 1.03 to 1.07) and PDD/DLB (HR 1.59 with 95% CI 1.14 to 2.20, reference: PD), after adjustment. Decreased risk of mortality was associated with female sex (HR 0.54, with a 95% CI 0.39 to 0.75), a pre-morbid CFS of <5 (HR 0.62, with a 95% CI 0.44 to 0.85, reference: 7-9), being vaccinated (HR 0.36, with a 95% CI 0.13 to 0.99) and having asymptomatic or mild respiratory COVID-19 symptoms (HR and 95% CI: 0.22, 0.12 to 0.39 and 0.27, 0.18 to 0.41, respectively, reference: respiratory support). No association was found after adjustment between mortality within 34-days and any other PD or COVID-19 features.

**Figure 5:**
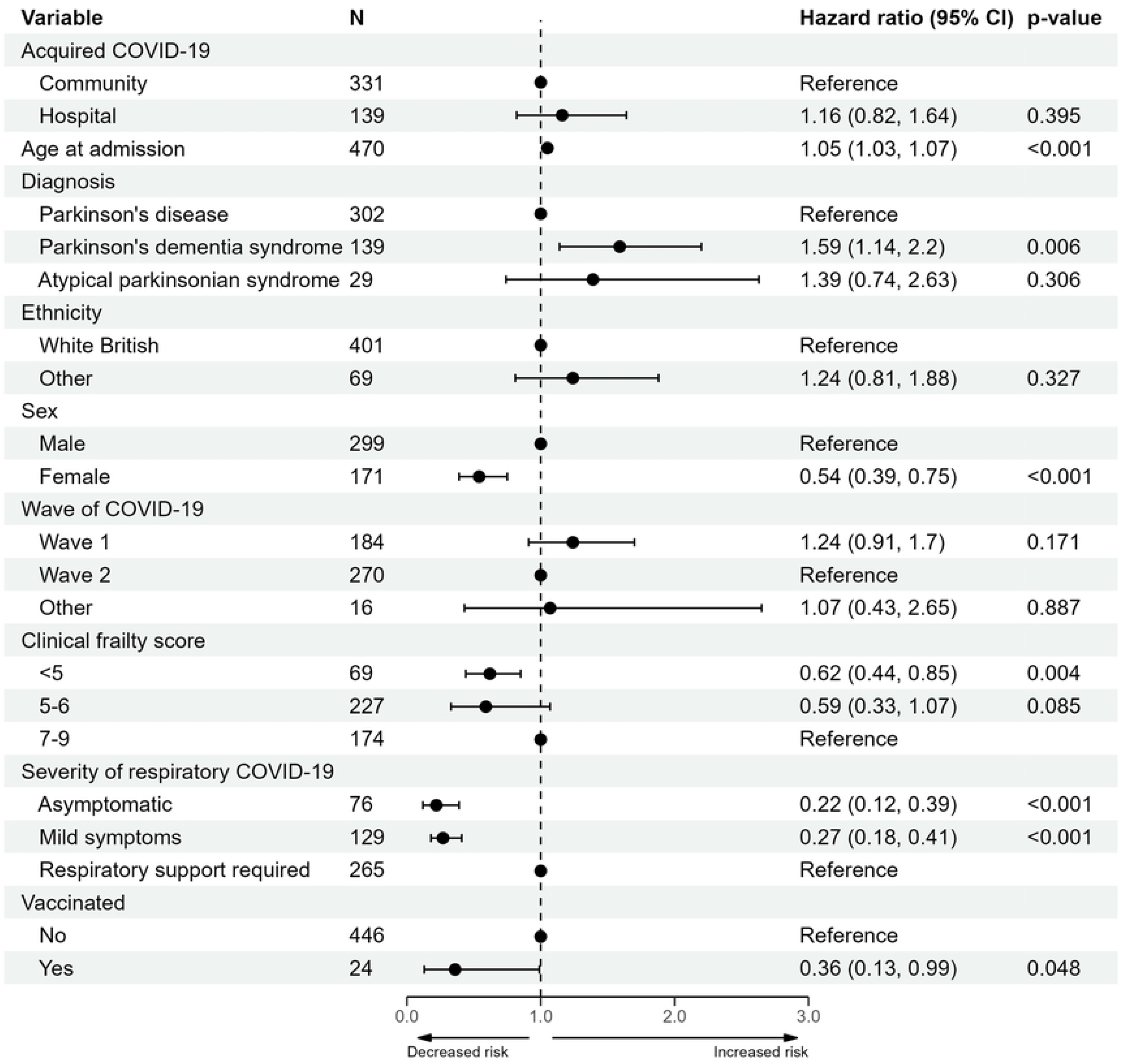
Forest plot of the multivariable Cox proportional hazards model of mortality within 34-days of a COVID-19 positive test. Abbreviations: confidence interval (CI).

As the date of the Sunday following death was captured during the study as a proxy for date of death, all patients included in the primary analysis have been censored at 34-days, with eight patients dying between days 28 and 34. We have conducted a sensitivity analysis investigating the effect of 28-day censoring; all sensitivity analyses found no substantial changes to the final mortality model (see Table S3.1, S3 Supporting information).

#### Requirement for respiratory support

Nine patients (1.6%) in the study were admitted to high-dependency care (HDU) or intensive care units (ICU). Over half of patients received respiratory support while in hospital (291/552, 52.7%) with the majority of these (266/291, 91%) receiving oxygen supplementation; 28.3% (156/552) had mild symptoms not requiring support and 19.0% (105/552) were asymptomatic from a respiratory perspective. Patients with asymptomatic and mild symptoms were combined to compare the odds of requiring respiratory support.

A forest plot presenting the results of the multivariable model is shown in Figure 6 (details of univariable and sensitivity analyses in Table S3.2, S3 Supporting information). Increased odds of requiring respiratory support were found to be associated with presence of a co-morbid chronic neurological disorder (odds ratio (OR) 1.81, with a 95% CI 1.06 to 3.09) and community-acquired COVID-19 (OR of hospital-acquired 0.54, with a 95% CI 0.36 to 0.81, reference: community) after adjustment. Sex, age, ethnicity and Parkinsonian syndrome diagnosis were not found to be associated with requiring respiratory support. Sensitivity analyses found no substantial changes to the multivariable model.

**Figure 6:**
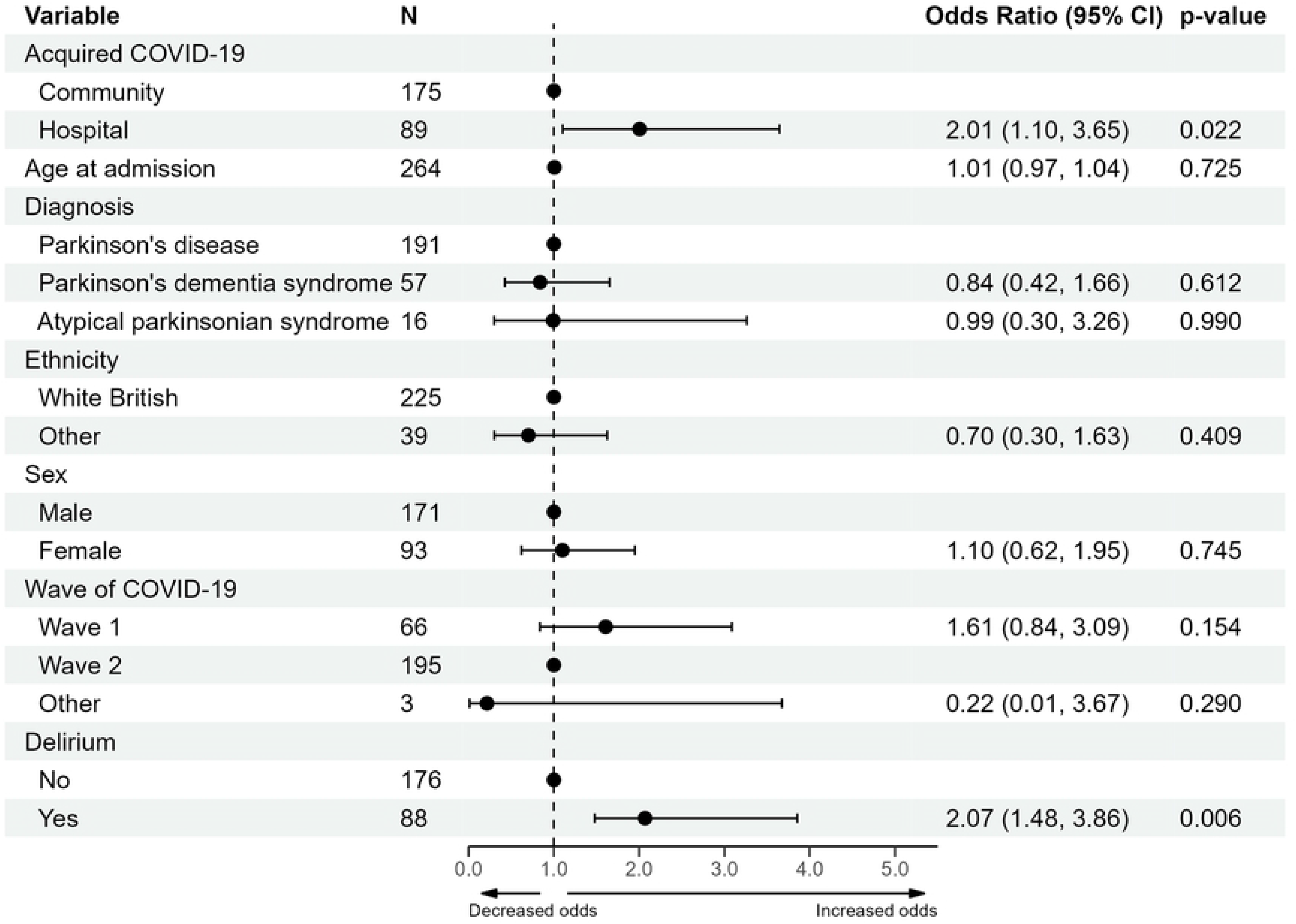
Forest plot of the multivariable logistic regression model of the requirement for respiratory support. Abbreviations: confidence interval (CI).

#### Increase in care requirement post-discharge

Increase in care was determined by change in location between admission and discharge, calculated for patients living at home prior to admission who were discharged from hospital. Of these 264 patients, 115 individuals (43.6%) had an increase in care and 149 (56.4%) remained at their pre-admission location at discharge. Results of the analysis are provided in Table S3.3 (see S3 Supporting information), while a forest plot of the multivariable results is provided in Figure 7. Increase in level of care post-discharge was associated with delirium associated with COVID-19 infection (OR 2.07, with a 95% CI 1.48 to 3.86) and hospital-acquired COVID-19 (OR 2.01, with a 95% CI 1.10 to 3.65).

**Figure 7:**
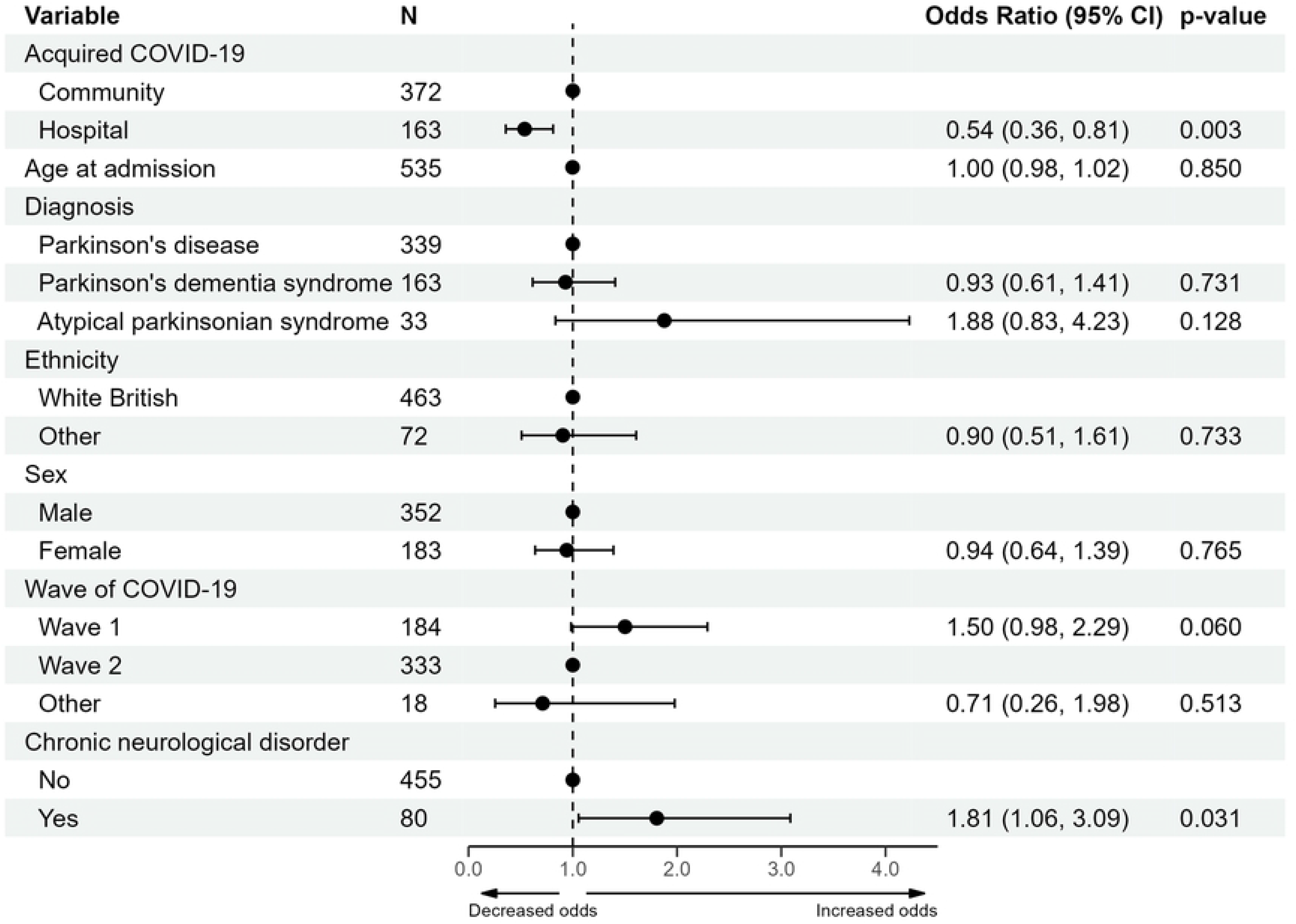
Forest plot of the multivariable logistic regression model of an increase in care post-discharge for patients admitted from their own home. Delirium is considered as delirium associated with COVID-19 infection. Abbreviations: confidence interval (CI).

#### Change in levodopa equivalent daily dose

Change in LEDD (mg) was calculated for the 359 individuals who were discharged and had both admission and discharge medication documented. There was no change in LEDD between admission and discharge for 76.0% of individuals (273/359), while 13.4% (48/359) had a decrease in LEDD and 10.6% (38/359) had an increase in LEDD. We observed no indication of an association between variables of clinical interest, diagnostic category and change in LEDD.

## Discussion

This is the first multicentre UK-based study to investigate demographics, comorbidities, clinical profiles and outcomes of PwP and APS who tested positive for COVID-19 in the hospital setting. The most common clinical presentations of COVID-19 in PwP and APS with community- and hospital-acquired COVID-19 were classic COVID-19 or respiratory symptoms and altered mental state. Comorbidities frequently reported were hypertension, dementia, chronic cardiac disease, and chronic kidney disease with an overall 34-day mortality rate of 38.4%. Increased age, male sex and diagnosis of PDD/DLB were associated with increased risk of mortality, while low pre-morbid CFS (<5), being vaccinated, and asymptomatic COVID-19 or mild respiratory symptoms were associated with reduced risk of mortality. Finally, while increased risk of requiring respiratory support was associated with presence of a co-morbid chronic neurological disorder and community-acquired COVID-19, patients with hospital-acquired COVID-19 and delirium required a higher level of care at discharge. Overall, our study extends previous findings on clinical features and outcomes of hospitalised PwP and APS with COVID-19.

In relation to demographics, our cohort of parkinsonian patients had a median age at admission of 80 years which was higher than one of the cohorts of hospitalised non-parkinsonian patients with COVID-19 (19). Of note, 73.4% (256/349) of our cohort of PD patients were over the age of 75 years compared to 56.3% of the estimated PD population in England (30). Our cohort included more male (66.1%) than female (33.9%) patients. Overall, these results are in line with known increased prevalence of PD in the elderly and in men (11, 12, 31, 32), as well as older age and male sex being risk factors for hospitalisation with COVID-19 generally (19, 33).

PD patients in our study were more frequently in the advanced motor phases of the disease (46% H&Y 4-5) than the general PD population (16.4% H&Y 4-5 (34)), although with similar prevalence of significant cognitive impairment (23.0% vs 24-31% in PD in the community (35)) and marked motor fluctuations (27.5% vs 29% in PD in the community (36)). In relation to frailty, which is known to be strongly associated with PD and mortality (37, 38), 78.3% of PD patients in our study were pre-morbidly frail (CFS ≥ 5), which is higher than a previously reported proportion of PwP with frailty in an ambulant community PwP population (33%) (39). Our finding is similar to previously reported levels of frailty in hospitalised PwP, where it has been found to predict mortality (40). Patients within our study were found to have an increased level of frailty compared to the reported cohorts of patients hospitalised with COVID-19 generally (41–43).

Irrespective of the modality of infection (hospital or community-acquired), the most common clinical presentations in our cohort of hospitalised parkinsonian patients with COVID-19 were classic COVID-19 symptoms including fever, shortness of breath, hyposmia and cough or respiratory symptoms in agreement with previous findings in the general population and in PwP (12, 19). However, we found 49.6% of community-acquired and 38.3% of hospital-acquired COVID-19 infections in our cohort also had altered mental state, with no respiratory symptoms in 3.9% and 9.6% respectively. Patients with altered mental state associated with COVID-19 were more likely to require increased care on discharge in our study. This aligns with findings from a single-centre study on hospitalised patients with COVID-19, where patients with delirium were less likely to be discharged home, and more frequently discharged to other hospitals, nursing homes or rehabilitation institutes (44). Inconsistent results on the contribution of delirium to mortality in COVID-19 have been published (45), and our data supports a lack of association. Although altered mental state is a common neurological manifestation of COVID-19 in hospitalised patients in general (up to 32%) (46–48), our data suggests that hospitalised PwP or APS and COVID-19 present more frequently with altered mental state than the general population, in line with previous findings and as expected in a population of patients with known central nervous system neurodegeneration (11). Cognitive impairment is frequently observed in PwP and APS and represents a well-known risk factor for development of delirium (49).

In our study, the most frequently reported comorbidities at admission were hypertension (46.3%), dementia (40.6%), chronic cardiac disease (35.8%), and chronic kidney disease (21.9%). These results only partly mirror those of the ISARIC study (19), which was based on UK hospitalised patients with COVID-19, where only 13.5% of patients were reported with dementia. This discrepancy might be due to the fact that cognitive impairment is an intrinsic feature of PD and APS as previously mentioned (49); in addition, our cohort included 170 patients with PDD/DLB who present with dementia as per diagnostic criteria (50). It is also worth mentioning that most PwP are non-smokers, which can partly explain the lower prevalence of chronic pulmonary disease in our cohort of patients than in the ISARIC study (51).

In relation to length of stay, the median time from positive test to discharge in our study was 12 and 16 days for patients with community- and hospital-acquired COVID-19, respectively, which is longer than data from other UK, US and Iranian cohorts (median hospital stay of 5, 10 and 7 days, respectively) (8, 11, 52) reflecting possible differences in study methodology, health care settings, spatial and temporal spread of the pandemic as well as intrinsic features of the study populations.

Our study showed an overall 34-day mortality rate of 38.4% which is in line with data deriving from other US and European cohorts of hospitalised PD patients with COVID-19 of 35.8% and 35.4%, respectively (12, 53). These mortality rates appear to be higher compared with data from the general population with COVID-19 hospitalised over the same period (overall mortality rate of 25%) (54), adding to the evidence that PwP have a significant risk of poor outcomes (5). Risk factors for increased mortality in our study were increased age, male sex and diagnosis of PD dementia while protective factors were low CFS (<5), being vaccinated, and COVID-19 with no or mild respiratory symptoms, in agreement with the literature on the general population and PD patients (3, 12, 19).

PwP/APS were more likely to require respiratory support if they had community-acquired COVID-19 or a co-morbid chronic neurological disorder. In our study, requirement for respiratory support was collected as a measure of severity of respiratory COVID-19. This might have unintentionally excluded individuals with worse respiratory COVID-19 than mild symptoms but did not have respiratory support available to them. Alternatively, there may have been individuals with mild symptoms that did not require respiratory support but received it as a precaution.

We found that patients with hospital-acquired COVID-19 and delirium had increased risk of a requiring a higher level of care at discharge (residential or nursing home or a local hospital). Development of delirium can signpost the presence of an underlying cognitive impairment which might become manifest during the infection and hospitalisation and, ultimately, have a detrimental effect on autonomy in performing activities of daily living (55).

Limitations of the study include the retrospective nature of the survey, perhaps contributing to the level of unknown or missing responses. Limited data on patients from diverse ethnicities were available and, therefore, we were unable to compare risks of poor COVID-19 outcomes between specific ethnic groups. The small number of APS patients limited our ability to evaluate outcomes within this group. COVID-19 testing policy in the UK changed during the course of the study and testing of asymptomatic patients and routine testing of hospital in-patients was uncommon during wave one; however, to account for the change in testing we have adjusted for wave in each model and conducted sensitivity analyses focused on wave two only, where we observed no substantial changes to any outcomes.

## Conclusion

In this first, multicentre, UK-based study we found that, compared with published data on COVID-19 admissions, PwP or APS hospitalised with COVID-19 have increased mortality and are more likely to require an increase in care level post-discharge. These findings may be attributable to an increased proportion of PwP/APS having dementia and cardiovascular co-morbidities, with COVID-19-associated delirium. PwP admitted with COVID-19 were older than community-dwelling PwP/APS, with more advanced Parkinson’s and higher pre-morbid frailty. Our study expands previous findings on clinical profiles and outcomes of hospitalised PwP and APS with COVID-19.

## Data Availability

Data will be made available on application to the sponsor with execution of an appropriate data sharing agreement.

## Acknowledgements

The views expressed are those of the authors and not necessarily those of the NHS, NIHR or Department of Health. The authors acknowledge the support of Parkinson’s UK who funded the study, the sponsor and the clinical and research teams who contributed to the study.

## Supporting information

**S1 Supporting information. Members of the COVID-19 PD UK study team.**

**S2 Supporting information. Tables of summary statistics.** Patient, Parkinson’s and COVID-19 characteristics, length of stay and a summary of outcomes.

**S3 Supporting information. Results of the regression analyses.** Univariable, multivariable and sensitivity analyses for the outcomes of mortality, requirement of respiratory support and increase in care.

## References

1. Wang C, Horby PW, Hayden FG, Gao GF. A novel coronavirus outbreak of global health concern. Lancet. 2020;395(10223):470–3.

2. Clift AK, Coupland CAC, Keogh RH, Diaz-Ordaz K, Williamson E, Harrison EM, et al. Living risk prediction algorithm (QCOVID) for risk of hospital admission and mortality from coronavirus 19 in adults: national derivation and validation cohort study. BMJ. 2020;371:m3731.

3. Hippisley-Cox J, Coupland CA, Mehta N, Keogh RH, Diaz-Ordaz K, Khunti K, et al. Risk prediction of covid-19 related death and hospital admission in adults after covid-19 vaccination: national prospective cohort study. BMJ. 2021;374.

4. Sulzer D, Antonini A, Leta V, Nordvig A, Smeyne RJ, Goldman JE, et al. COVID-19 and possible links with Parkinson’s disease and parkinsonism: from bench to bedside. NPJ Parkinsons Dis. 2020;6:18.

5. Fearon C, Fasano A. Prevalence and outcomes of Covid-19 in Parkinson’s disease: Acute settings and hospital. Int Rev Neurobiol. 2022;165:35–62.

6. Antonini A, Leta V, Teo J, Chaudhuri KR. Outcome of Parkinson’s Disease Patients Affected by COVID-19. Mov Disord. 2020;35(6):905–8.

7. Fasano A, Elia AE, Dallocchio C, Canesi M, Alimonti D, Sorbera C, et al. Predictors of COVID-19 outcome in Parkinson’s disease. Parkinsonism Relat Disord. 2020;78:134–7.

8. Fathi M, Taghizadeh F, Mojtahedi H, Zargar Balaye Jame S, Markazi Moghaddam N. The effects of Alzheimer’s and Parkinson’s disease on 28-day mortality of COVID-19. Rev Neurol (Paris). 2022;178(1-2):129–36.

9. Hainque E, Grabli D. Rapid worsening in Parkinson’s disease may hide COVID-19 infection. Parkinsonism Relat Disord. 2020;75:126–7.

10. Kobylecki C, Jones T, Lim CK, Miller C, Thomson AM. Phenomenology and Outcomes of In-Patients With Parkinson’s Disease During the Coronavirus Disease 2019 Pandemic. Mov Disord. 2020;35(8):1295–6.

11. Nwabuobi L, Zhang C, Henchcliffe C, Shah H, Sarva H, Lee A, et al. Characteristics and Outcomes of Parkinson’s Disease Individuals Hospitalized with COVID-19 in a New York City Hospital System. Mov Disord Clin Pract. 2021;8(7):1100–6.

12. Parihar R, Ferastraoaru V, Galanopoulou AS, Geyer HL, Kaufman DM. Outcome of Hospitalized Parkinson’s Disease Patients with and without COVID-19. Mov Disord Clin Pract. 2021;8(6):859–67.

13. Sainz-Amo R, Baena-Álvarez B, Pareés I, Sánchez-Díez G, Pérez-Torre P, López-Sendón JL, et al. COVID-19 in Parkinson’s disease: what holds the key? J Neurol. 2021;268(8):2666–70.

14. Salari M, Zali A, Ashrafi F, Etemadifar M, Sharma S, Hajizadeh N, et al. Incidence of Anxiety in Parkinson’s Disease During the Coronavirus Disease (COVID-19) Pandemic. Mov Disord. 2020;35(7):1095–6.

15. Vignatelli L, Zenesini C, Belotti LMB, Baldin E, Bonavina G, Calandra-Buonaura G, et al. Risk of Hospitalization and Death for COVID-19 in People with Parkinson’s Disease or Parkinsonism. Mov Disord. 2021;36(1):1–10.

16. Xu Y, Surface M, Chan AK, Halpern J, Vanegas-Arroyave N, Ford B, et al. COVID-19 manifestations in people with Parkinson’s disease: a USA cohort. J Neurol. 2022;269(3):1107–13.

17. Zhai H, Lv Y, Xu Y, Wu Y, Zeng W, Wang T, et al. Characteristic of Parkinson’s disease with severe COVID-19: a study of 10 cases from Wuhan. J Neural Transm (Vienna). 2021;128(1):37–48.

18. Zhang Q, Schultz JL, Aldridge GM, Simmering JE, Narayanan NS. Coronavirus Disease 2019 Case Fatality and Parkinson’s Disease. Mov Disord. 2020;35(11):1914–5.

19. Docherty AB, Harrison EM, Green CA, Hardwick HE, Pius R, Norman L, et al. Features of 20 133 UK patients in hospital with covid-19 using the ISARIC WHO Clinical Characterisation Protocol: prospective observational cohort study. BMJ. 2020;369:m1985.

20. Rockwood K, Song X, MacKnight C, Bergman H, Hogan DB, McDowell I, et al. A global clinical measure of fitness and frailty in elderly people. Cmaj. 2005;173(5):489–95.

21. The ABN Executive in association with subspecialist Advisory Groups. Association of British Neurologists Guidance on COVID-19 for people with neurological conditions, their doctors and carers. 2020 9 April 2020.

22. Office for National Statistics. Coronavirus (COVID-19) latest insights: Comparisons 2022 [Available from: https://www.ons.gov.uk/peoplepopulationandcommunity/healthandsocialcare/conditionsanddiseases/articles/coronaviruscovid19latestinsights/overview.

23. Office for National Statistics. Coronavirus (COVID-19) latest insights: Infections 2022 [Available from: https://www.ons.gov.uk/peoplepopulationandcommunity/healthandsocialcare/conditionsanddiseases/articles/coronaviruscovid19latestinsights/infections.

24. R Development Core Team. R: A language and environment for statistical computing. Vienna, Austria: R Foundation for Statistical Computing; 2022.

25. Therneau T. A package for Survival Analysis in R. 3.2-13 ed2021.

26. Brooks ME, Kristensen K, van Benthem KJ, Magnusson A, Berg CW, Nielsen A, et al. glmmTMB Balances Speed and Flexibility Among Packages for Zero-inflated Generalized Linear Mixed Modeling. The R Journal. 2017;9(2):378–400.

27. Boyes R. Forester: An R package for creating publication-ready forest plots 0.3.0 ed2021.

28. UK Health Security Agency. Coronavirus (COVID-19) in the UK 2022 [Available from: https://coronavirus.data.gov.uk/details/healthcare.

29. Nuffield Department of Population Health. RECOVERY Trial 2023 [Available from: https://www.recoverytrial.net/.

30. Parkinson’s UK. The prevalence and incidence of Parkinson’s in the UK. 2017.

31. Van Den Eeden SK, Tanner CM, Bernstein AL, Fross RD, Leimpeter A, Bloch DA, et al. Incidence of Parkinson’s disease: variation by age, gender, and race/ethnicity. Am J Epidemiol. 2003;157(11):1015–22.

32. Scherbaum R, Kwon EH, Richter D, Bartig D, Gold R, Krogias C, et al. Clinical Profiles and Mortality of COVID-19 Inpatients with Parkinson’s Disease in Germany. Mov Disord. 2021;36(5):1049–57.

33. Chaturvedi R, Lui B, Aaronson JA, White RS, Samuels JD. COVID-19 complications in males and females: recent developments. J Comp Eff Res. 2022;11(9):689–98.

34. Zhao YJ, Wee HL, Chan YH, Seah SH, Au WL, Lau PN, et al. Progression of Parkinson’s disease as evaluated by Hoehn and Yahr stage transition times. Mov Disord. 2010;25(6):710–6.

35. Aarsland D, Zaccai J, Brayne C. A systematic review of prevalence studies of dementia in Parkinson’s disease. Mov Disord. 2005;20(10):1255–63.

36. Schrag A, Quinn N. Dyskinesias and motor fluctuations in Parkinson’s disease. A community-based study. Brain. 2000;123 (Pt 11):2297–305.

37. Blomaard LC, van der Linden CMJ, van der Bol JM, Jansen SWM, Polinder-Bos HA, Willems HC, et al. Frailty is associated with in-hospital mortality in older hospitalised COVID-19 patients in the Netherlands: the COVID-OLD study. Age Ageing. 2021;50(3):631–40.

38. McMillan JM, Michalchuk Q, Goodarzi Z. Frailty in Parkinson’s disease: A systematic review and meta-analysis. Clin Park Relat Disord. 2021;4:100095.

39. Ahmed NN, Sherman SJ, Vanwyck D. Frailty in Parkinson’s disease and its clinical implications. Parkinsonism Relat Disord. 2008;14(4):334–7.

40. Torsney KM, Romero-Ortuno R. The Clinical Frailty Scale predicts inpatient mortality in older hospitalised patients with idiopathic Parkinson’s disease. J R Coll Physicians Edinb. 2018;48(2):103–7.

41. Martí-Pastor A, Moreno-Perez O, Lobato-Martínez E, Valero-Sempere F, Amo-Lozano A, Martínez-García M, et al. Association between Clinical Frailty Scale (CFS) and clinical presentation and outcomes in older inpatients with COVID-19. BMC Geriatr. 2023;23(1):1.

42. Andrés-Esteban EM, Quintana-Diaz M, Ramírez-Cervantes KL, Benayas-Peña I, Silva-Obregón A, Magallón-Botaya R, et al. Outcomes of hospitalized patients with COVID-19 according to level of frailty. PeerJ. 2021;9:e11260.

43. Aw D, Woodrow L, Ogliari G, Harwood R. Association of frailty with mortality in older inpatients with Covid-19: a cohort study. Age Ageing. 2020;49(6):915–22.

44. Wilke V, Sulyok M, Stefanou MI, Richter V, Bender B, Ernemann U, et al. Delirium in hospitalized COVID-19 patients: Predictors and implications for patient outcome. PLoS One. 2022;17(12):e0278214.

45. White L, Jackson T. Delirium and COVID-19: a narrative review of emerging evidence. Anaesthesia. 2022;77 Suppl 1:49–58.

46. Liotta EM, Batra A, Clark JR, Shlobin NA, Hoffman SC, Orban ZS, et al. Frequent neurologic manifestations and encephalopathy-associated morbidity in Covid-19 patients. Ann Clin Transl Neurol. 2020;7(11):2221–30.

47. Attia AS, Hussein M, Aboueisha MA, Omar M, Youssef MR, Mankowski N, et al. Altered mental status is a predictor of poor outcomes in COVID-19 patients: A cohort study. PLoS One. 2021;16(10):e0258095.

48. Kenerly MJ, Shah P, Patel H, Racine R, Jani Y, Owens C, et al. Altered mental status is an independent predictor of mortality in hospitalized COVID-19 patients. Ir J Med Sci. 2022;191(1):21–6.

49. Aarsland D, Batzu L, Halliday GM, Geurtsen GJ, Ballard C, Ray Chaudhuri K, et al. Parkinson disease-associated cognitive impairment. Nat Rev Dis Primers. 2021;7(1):47.

50. Emre M, Aarsland D, Brown R, Burn DJ, Duyckaerts C, Mizuno Y, et al. Clinical diagnostic criteria for dementia associated with Parkinson’s disease. Mov Disord. 2007;22(12):1689–707; quiz 837.

51. Ritz B, Lee PC, Lassen CF, Arah OA. Parkinson disease and smoking revisited: ease of quitting is an early sign of the disease. Neurology. 2014;83(16):1396–402.

52. Ellis PK, Davies ML, Gray WK, Barber M, Bolnykh I, Sadler M, et al. The Cause and Duration of Emergency Admissions to Hospital in People with Idiopathic Parkinson’s Disease, Under the Care of a UK Service, During the First Year of the COVID-19 Pandemic. J Parkinsons Dis. 2022;12(6):1833–40.

53. Scherbaum R, Kwon EH, Richter D, Bartig D, Gold R, Krogias C, et al. Clinical Profiles and Mortality of COVID-19 Inpatients with Parkinson’s Disease in Germany. Movement Disorders. 2021;36(5):1049–57.

54. Gray WK, Navaratnam AV, Day J, Wendon J, Briggs TWR. COVID-19 hospital activity and in-hospital mortality during the first and second waves of the pandemic in England: an observational study. Thorax. 2022;77(11):1113–20.

55. Green S, Perrott SL, McCleary A, Sleeman I, Maple-Grødem J, Counsell CE, et al. First delirium episode in Parkinson’s disease and parkinsonism: incidence, predictors, and outcomes. NPJ Parkinsons Dis. 2021;7(1):92.

